# Estimation of excess all-cause mortality due to COVID-19 in Thailand

**DOI:** 10.1101/2022.01.07.22268886

**Authors:** Chaiwat Wilasang, Charin Modchang, Thanchanok Lincharoen, Sudarat Chadsuthi

## Abstract

Thailand has experienced the most prominent COVID-19 outbreak, resulting in a new record for COVID-19 cases and deaths in 2021. To assess the influence of the COVID-19 outbreak on mortality, we estimated excess all-cause and pneumonia mortality in Thailand during the COVID-19 outbreak from April to October 2021. We used the previous five years’ mortality to estimate the baseline number of deaths using generalized linear mixed models (GLMMs). The models were adjusted for seasonality and demographics. We found that the estimated cumulative excess death was 14.3% (95% CI: 8.6%-18.8%) higher than the baseline. The results also showed that the excess deaths in males were higher than in females by approximately 26.3%. The excess deaths directly caused by the COVID-19 infections accounted for approximately 75.0% of the all-cause excess deaths. Furthermore, excess pneumonia deaths were also found to be 26.2% (95% CI: 4.8%-46.0%) above baseline. There was a significant rise in excess fatalities, especially in the older age groups. Therefore, the age and sex structure of the population are essential to assessing the mortality impact of COVID-19. Our modeling results could potentially provide insights into the COVID-19 outbreaks and provide a guide for outbreak control and intervention.

## Introduction

Thailand has recently experienced the most prominent coronavirus disease 2019 (COVID-19) outbreak in 2021 (1, 2). To mitigate the disease transmission, the Thai government implemented social distancing and stringent lockdown measures throughout the country and imposed international travel restrictions and nighttime curfews (3). Even though the interventions have proven to be effective in slowing down the spread of the disease, during the peak of the epidemic wave, hospitals in Thailand were running out of beds to accommodate COVID-19 patients and patients of other diseases (4-6). Although the Thai government had set up the field hospitals (7) for treating COVID-19 patients, the disease’s toll on mortality was still high (8). According to a report by the Department of Disease Control, Ministry of Public Health, Thailand^3^, there were 19,111 officially reported COVID-19 deaths since the beginning to October 31, 2021. This number of deaths reflects the significant impact of the COVID-19 outbreaks on mortality in Thailand.

To assess the impact of the outbreak on mortality, the approach for estimating excess mortality was utilized to quantify the increased deaths compared with the expected mortality. Excess mortality estimation has previously been used to investigate the effects of pollution (9), climate change (10), and epidemics (e.g., influenza (11, 12), chikungunya (13), and HIV (14)). Moreover, previous studies have also applied this approach to COVID-19, focusing on estimating the mortality burden of the COVID-19 outbreak using statistical models. In general, statistical models usually incorporate risk factors related to death (e.g., sex, age group, time, seasonality, and demographic change) as indicator functions for excess mortality estimation. For instance, J. M. Aburto et al., (15) estimated the number of age- and sex-specific excess deaths adjusted for seasonality using generalized additive models (GAM).

Estimating age- and sex-specific mortality is an essential consideration in COVID-19-related death. Age-specific mortality may vary by age due to a wide variety of factors. Most fatalities from COVID-19 have been reported to be relatively high in older age groups. Recent studies reveal that the proportion of elderly deaths has dramatically increased in many countries (15-18). Thus, disregarding the age structure of the population may result in inaccurate estimates of excess mortality. Sex-specific mortality is also important in estimating excess mortality. Different patterns of COVID-19 mortality have been observed among males and females in many countries (15, 17-19). Therefore, tracking age-and sex-specific deaths is an important component of health surveillance.

In this study, we aimed to estimate excess mortality in Thailand from April to October 2021 to assess the impact of the COVID-19 outbreaks on mortality. We estimated the baseline number of deaths in a typical year (2015-2019) without COVID-19 using a generalized linear mixed model (GLMM). Our model incorporated several factors such as sex, age group, time, seasonality, and demographics. We then estimated the all-cause mortality from April to October 2021 compared to baseline.

## Materials and methods

### Data

We collected the monthly all-cause mortality data in Thailand from Official Statistics Registration Systems in the human mortality database (20). The database compiles mortality statistics from Thailand’s Bureau of Registration Administration. The information was presented in the form of tables with age and gender categories. The mortality data from 2015 to 2019 was utilized to estimate the baseline mortality in the absence of COVID-19 in 2020-2021. The number of deaths was categorized into six age groups (0-14, 15-18, 19-24, 25-34, 35-60, and over 60 years of age).

The number of daily COVID-19 cases and deaths in Thailand were retrieved from the Center for Systems Science and Engineering (CSSE) at Johns Hopkins University (21). Daily COVID-19 mortality data were summarized into monthly data for consistency with the monthly all-cause mortality. According to the COVID-19 mortality data, the number of confirmed deaths started to increase rapidly in early April 2021 (see **Fig S1** in the supplementary information). We, therefore, focused our attention on estimating the excess deaths from April to October 2021, a period in which the cumulative number of COVID-19 deaths exceeds 100.

The monthly pneumonia mortality data were obtained from the Bureau of Epidemiology, Department of Disease Control, MoPH, Thailand (22). The data was country-level without age and sex stratification, as that information was not available. We used the pneumonia mortality data from 2015 to 2019 to estimate the pneumonia baseline mortality in the absence of COVID-19 in 2020-2021.

We also obtained yearly population estimates from the Population Pyramids of the World from 2015 to 2021 (23). The single-year population counts were aggregated to match the gender and age group of the mortality data. We employed the standard interpolation approach (15) to estimate the monthly population by gender and age group. The monthly population counts were used as an offset in the modelling strategy to calculate the baseline mortality in Thailand.

### Estimating the excess mortality

In order to estimate the excess mortality, we started our analysis by investigating the distribution of the observed sex-specific death counts from 2015 to 2019. We then used mortality data from January 2015 to December 2019 to estimate the baseline mortality in the absence of COVID-19. The mortality data were fitted by generalized linear mixed models (GLMMs) with a negative binomial link function. The models included linear mortality trends by gender, age, and seasonality. The structure of the models was as follows:

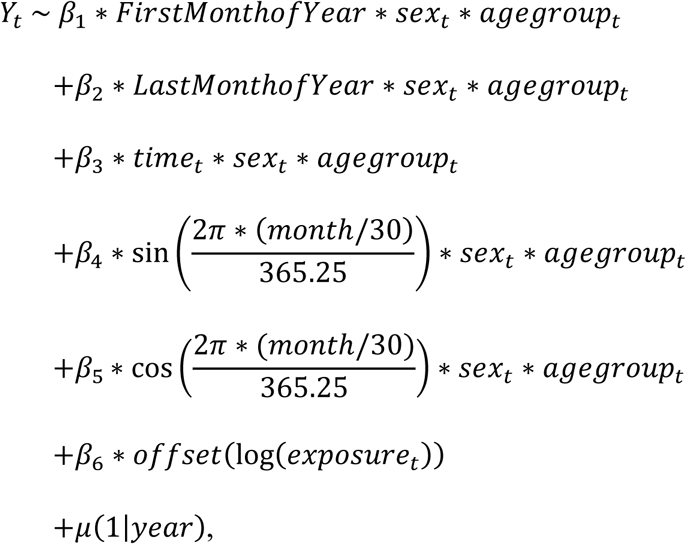

where *Y*_*t;*_ is the expected number of deaths at month *t. sex*_*t*_and *agegroup*_*t*_ represent the gender and age group at month *t. FirstMonthofYear* and *LastMonthofYear* are indicator variables (1: yes; 0: no). *time*_*t*_ indicates the month starting from January 2015 to December 2019. Furthermore, the featuring trigonometric terms represent the seasonal effect, and all the terms are fully interacted with gender and age group. *month* indicates the month of the year. This model also captures a yearly trend of changing population structure. The monthly average population by age group and gender at month *t* (*exposure*_*t*_) was used as an offset in the modelling. Here, *year* is a random effect in the model. The baseline was then created by averaging the death counts observed in each month starting from January 2015 to December 2019. We also compare our model with other GLMMs. We investigated how adjustments for seasonality might affect the baseline estimation. We constructed 4 models with 2 patterns of seasonality, including a full year period and a half-year period of seasonality (for full details, see the supplementary information).

We then projected the baseline mortality forward until October 2021. The excess mortality in each month was defined as the number of reported deaths minus the baseline prediction. We subsequently summed the excess mortality estimates across all months, starting from April 2021 to October 2021, yielding the cumulative estimate of the excess mortality in Thailand.

All the analyses were implemented in R statistical software version 3.6.3. To calculate the 95% predictive intervals (PI), we sampled death counts from a negative binomial distribution, following an approach developed by J. M. Aburto et al., (15).

## Results

Thailand has been plagued by competition between Alpha and Delta variants since April 2021 (1, 2), resulting in Thailand’s most ever severe epidemic wave. The number of COVID-19 deaths has increased across the country (**Fig. S1**). Until August 2021, daily deaths reached their peak with an average of 235 deaths per day (24). Following that, the death toll continued to decrease, with an average of 99 deaths per day in October (25). During April to late August, there were more than 10,000 officially COVID-19 confirmed deaths in Thailand.

This study estimated excess mortality in Thailand during the COVID-19 outbreak using GLMM. To do so, we first evaluated the ability of different models to retrospectively explain the mortality data from 2015 to 2019. The baseline mortality estimated using different models described in the method section is shown in **Fig. S2-S9** in the supplement information. A comparison of the models showed that GLMMs with a full year of seasonality were more consistent with the data. We found that the GLMM with age- and sex-specific effects for special months (i.e., the first and last months of a year) yields the best result based on the Akaike information criterion (AIC) (**Table S1**. in supplement information). The observed number of deaths is indicated by the points, and the expected number of deaths is illustrated by the solid line along with their 95% CI. Overall, the trends of monthly deaths generated from the model agree with the reported data. We also predicted the baseline mortality in 2020-2021 based on the 2015-2019 mortality data. We found that the predicted baseline in 2020-2021 during the COVID-19 outbreak was higher than the reported mortality data in the younger age group (0-14) of men, whereas the predicted baseline in other age groups agreed with the mortality data.

### All-cause mortality during 2015 – 2021 in Thailand

All-cause deaths by age group and sex from 2015 to 2021 in Thailand are illustrated in **Fig. 1** and **2**. To compare all-cause mortality trends in each age group, the number of reported total all-cause deaths was divided by the population in each age group. We observed some annual seasonal patterns of all-cause deaths, with higher death rates in the first and last months of a year for both men and women. The changes in mortality rate that occurred during the COVID-19 outbreak varied by age group. The mortality rate continually declines in the younger age group (0-24) for both men and women. However, in age groups older than 25 years, the mortality was characteristic in which the unusual mortality peak was observed. The mortality rate rapidly rises from June to August, with the mortality rate among men being higher than women. In the age group 36-60, the mortality rate was approximately 2-fold higher among men than women, and 1.5-fold in the age group over 60 years of age.

**Figure 1.**
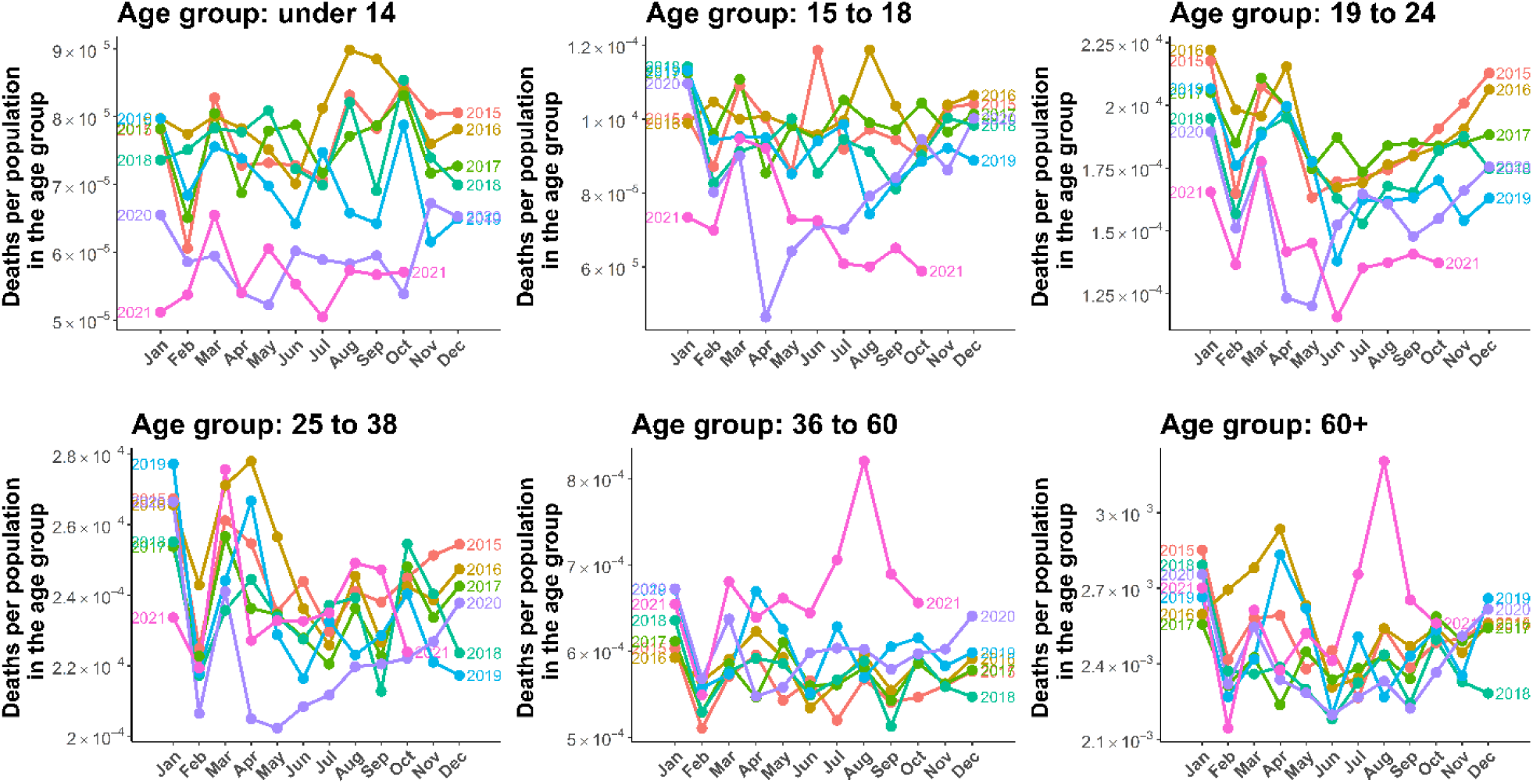
Men all-cause mortality. Lines illustrate the mortality by six age groups (0-14, 15-18, 19-24, 25-34, 35-60, and over 60 years of age) starting from January 2015 to October 2021.

**Figure 2.**
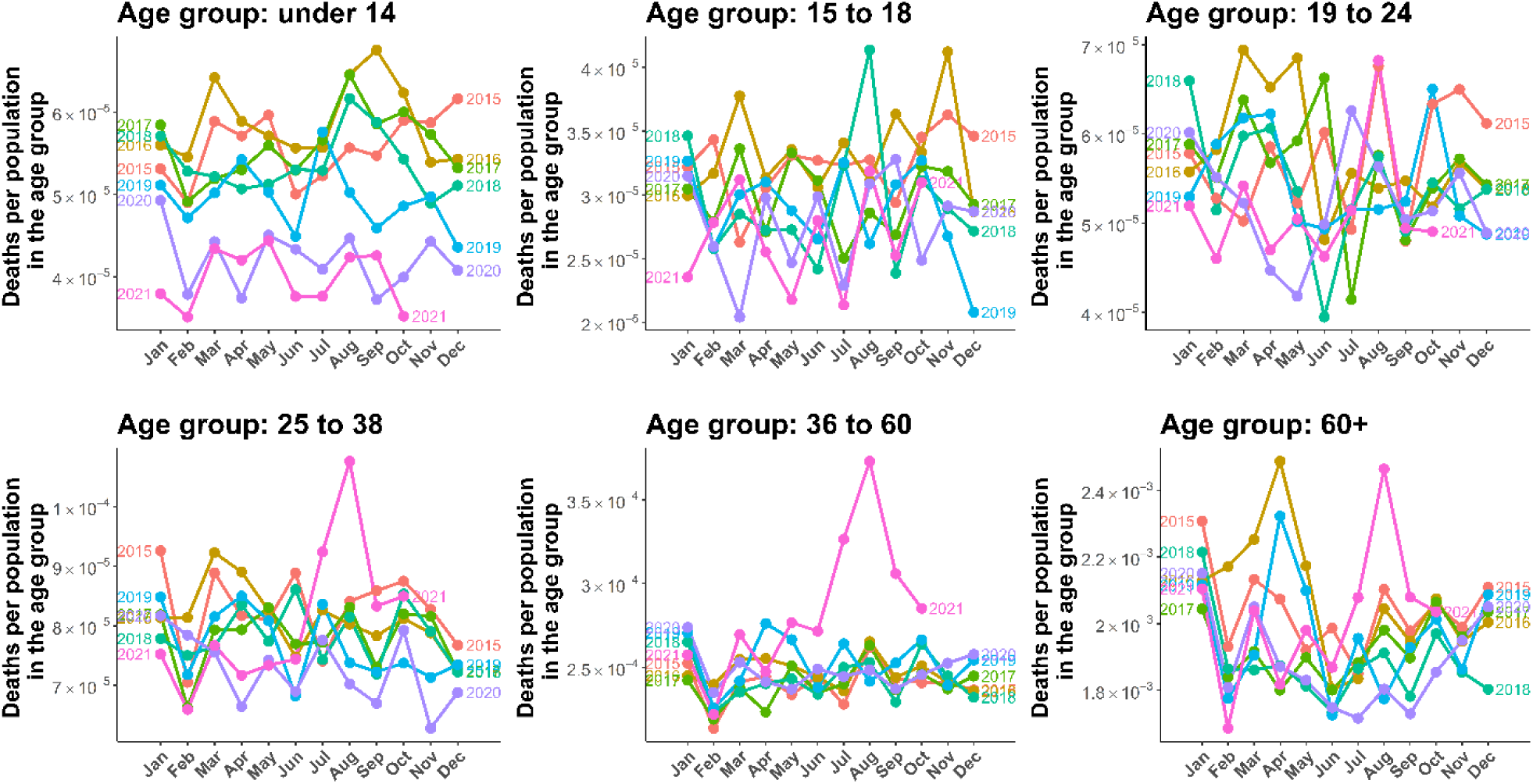
Women all-cause mortality. Lines illustrate the mortality by six age groups (0-14, 15-18, 19-24, 25-34, 35-60, and over 60 years of age) starting from January 2015 to October 2021.

### Cumulative excess mortality

The estimated cumulative excess deaths from April to October 2021 for men and women is shown in **Fig. 3**. The expected number of all-cause excess deaths up to October 31, 2021 was 25,486 (95% CI: 13,913-36,862). We found that the cumulative death counts had risen by 14.3% (95% CI: 8.6%-18.8%) above the baseline. The results showed that male excess deaths accounted for 55.8% (14,223) of total excess deaths, while female excess deaths accounted for 44.2% (11,263). These excess deaths included deaths caused by COVID-19 and other causes. Up to October 31, 2021, there were 19,111 officially reported COVID-19 deaths. After classifying the deaths caused by COVID-19, we found that the deaths attributed to COVID-19 accounted for 75.0% of the excess deaths throughout the study period, with the remaining 25.0% (6,375) being indirectly attributed to COVID-19.

**Figure 3.**
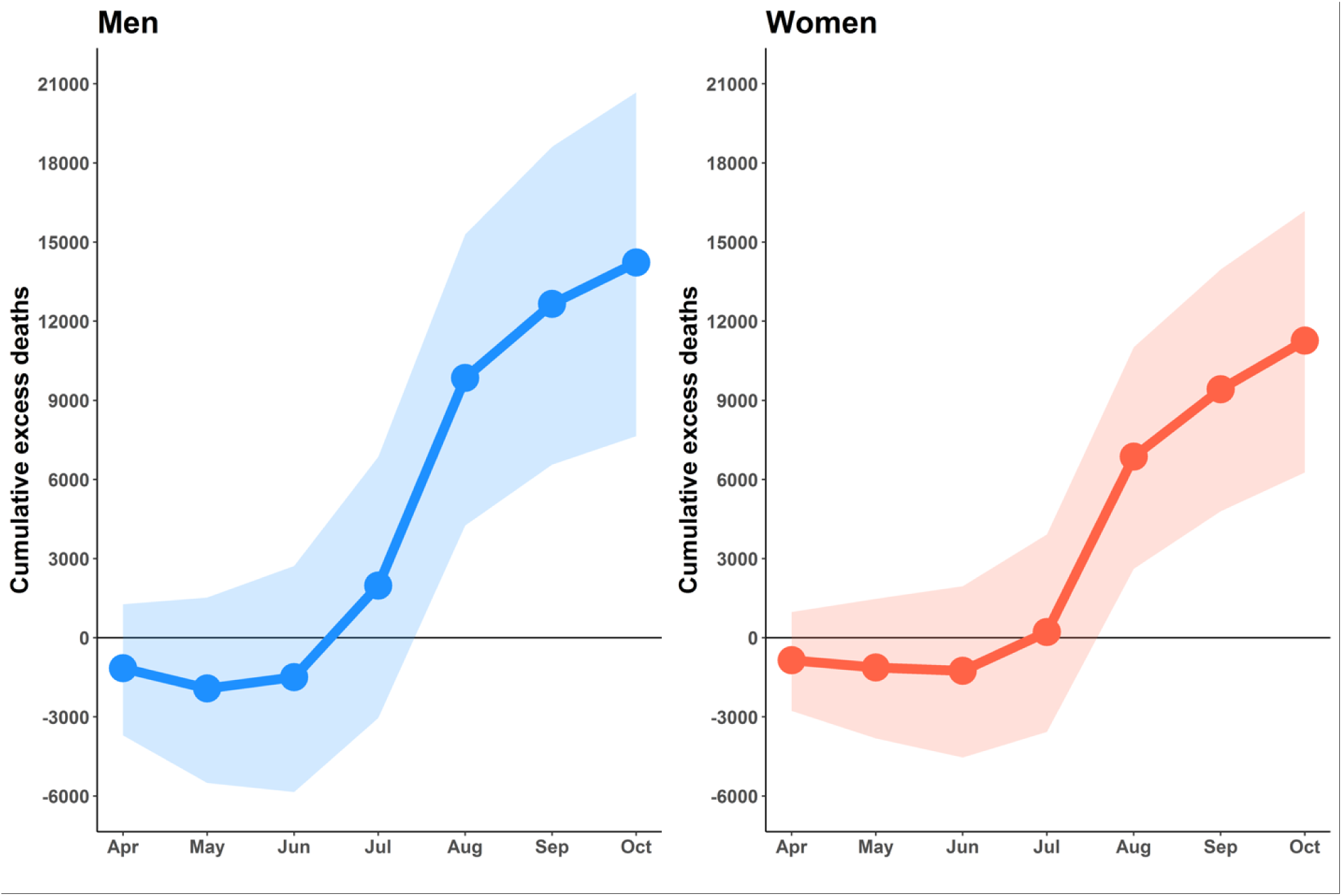
Cumulative excess deaths by gender starting from April to October 2021. Lines illustrate the cumulative excess deaths by gender. Shaded areas show 95% prediction intervals.

According to the time series of all-cause excess deaths, the number of excess deaths began to rise above the baseline in July 2021 (Fig. 3), which was the first month that the daily number of COVID-19 deaths exceeds a hundred. For both men and women, excess deaths were obviously observed in age groups older than 25 years, whereas excess in age groups 15-18 and 19-24 were below the baseline with -18.1% and -12.0%, respectively (**Fig. 4**). For children aged 0-14, the excess deaths decreased -19.6% compared with the expected level. The increases in mortality were observed in people aged 25-34 and 35-60 years, with 5.9% and 8.4% more deaths than expected. Among the six age groups, the oldest age groups (> 60 years of age) were most affected by the COVID-19 outbreak. People in the age group of more than 60 years were exposed to death at an 11.6% higher than baseline, with men and women having roughly similar trend.

**Figure 4.**
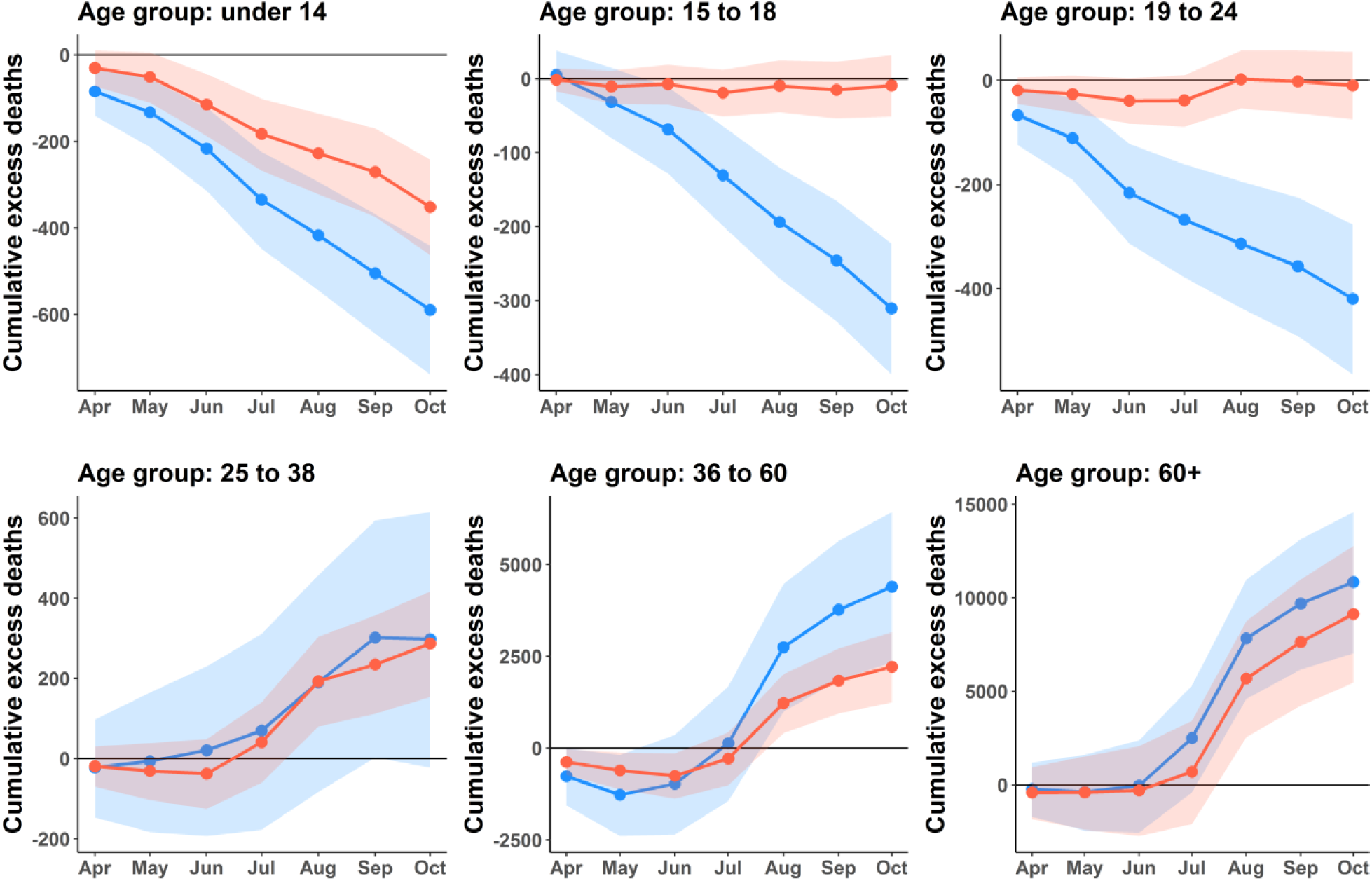
Cumulative excess deaths by gender and age groups. Lines illustrate the cumulative excess deaths by gender and age groups. Blue and orange lines represent men and women, respectively. Shaded areas indicate the 95% prediction intervals.

### Correlation of excess deaths and COVID-19

Our study demonstrated that the excess mortality across the country had increased as a consequence of the COVID-19 epidemic. The results revealed that the excess deaths substantially increased over the course of the outbreak (**Fig. 5**). Our model predicted that the excess mortality counts began to rise after the outbreak in April 2021. The disease spread to many provinces, with a maximum of 23,021 newly confirmed cases per day on August 13, 2021 (25). We found that the trend of all-cause excess mortality agrees with the COVID-19 confirmed cases observed by the surveillance system. Quantitatively, the Pearson correlation coefficient between the cumulative cases and cumulative excess deaths was found to be 0.9912 (95% CI: 0.9392-0.9987). This correlation suggests that the COVID-19 outbreaks had an impact on excess mortality.

**Figure 5.**
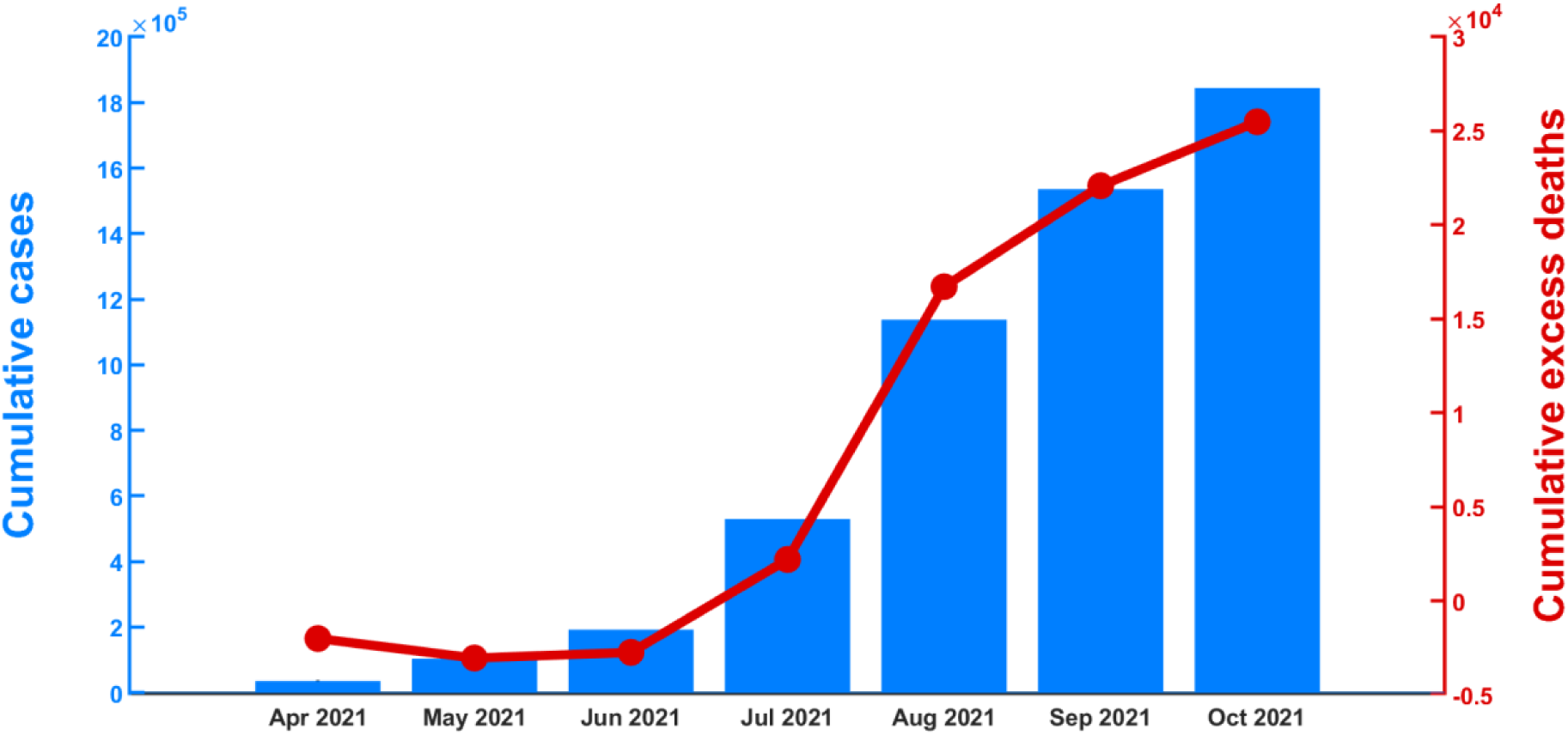
Excess deaths and confirmed COVID-19 cases. Bars show cumulative COVID-19 cases from April to October 2021. The red line indicates cumulative all-cause excess deaths.

### Pneumonia excess mortality

In this study, we also estimated the pneumonia excess mortality during the COVID-19 outbreak (for full details, see the supplementary information). We found that the trend of pneumonia deaths continually decreased from 2015 to 2020 (**Fig. S10** in the supplementary information). However, we observed unexpected pneumonia deaths in May 2021. Using the GLMMs model, we estimated pneumonia excess deaths between April and October 2021. The pneumonia excess deaths are illustrated in **Fig 6**. The results suggested that the number of pneumonia excess deaths exceeded the baseline by 26.2% (95% CI: 4.8%-46.0%) as of October 31, 2021. The cumulative pneumonia excess deaths were found to be 34 (95% CI: 6-58). Furthermore, the results showed that pneumonia excess deaths began to rise rapidly in April 2021, the same time period that COVID-19 began to spread in Thailand and decreased slowly afterward.

**Figure 6.**
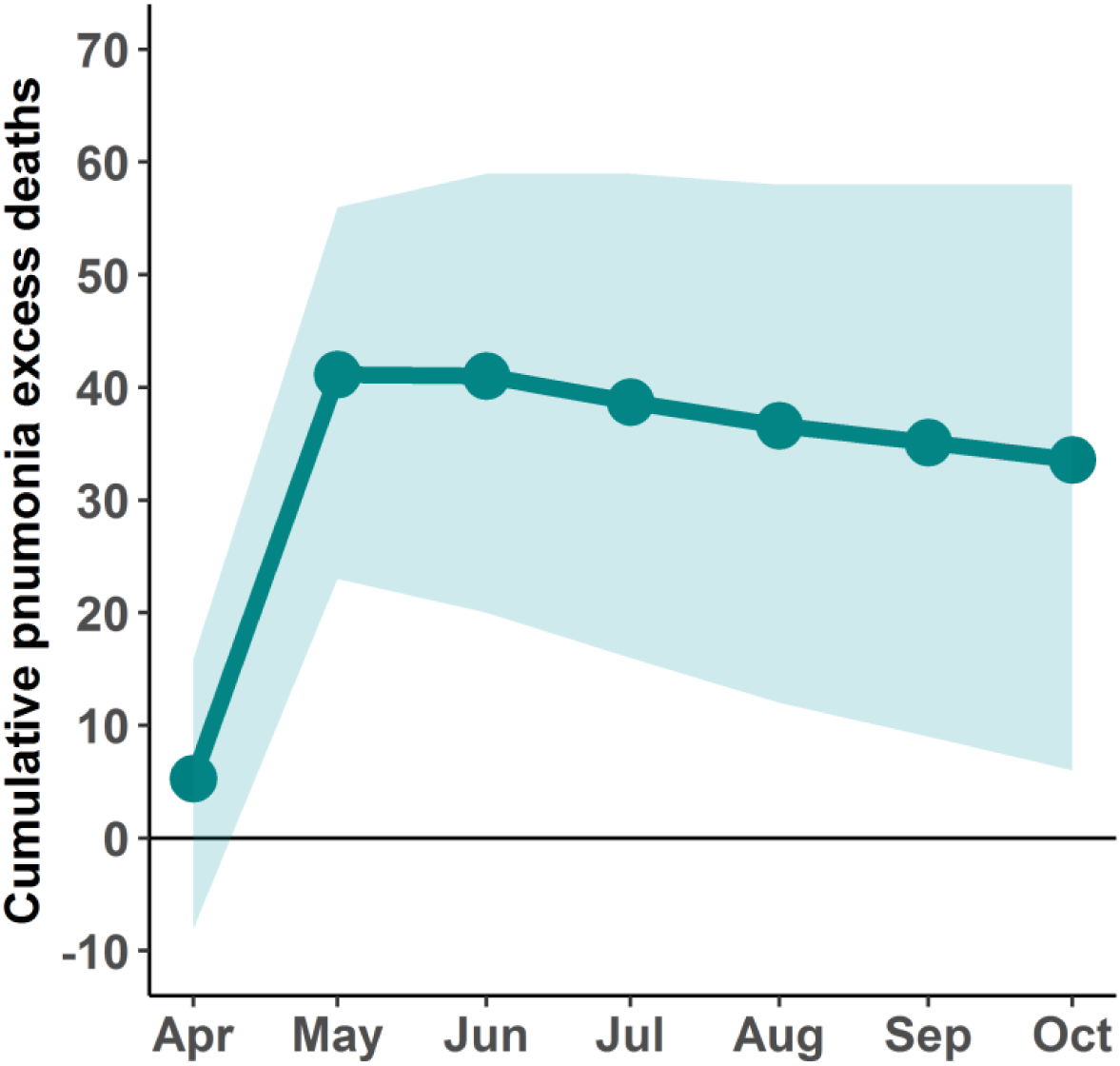
Cumulative pneumonia excess deaths. Line illustrates the cumulative Pneumonia excess deaths starting from April to October 2021, and the shaded area indicates the 95% CI.

## Discussion

In this study, we investigated the impact of COVID-19 outbreaks on excess mortality in Thailand. We estimated all-cause excess mortality in 2021 based on historical mortality data from 2015 to 2019. We then analyzed the age- and sex-specific mortality during 2020-2021.

Using our best model, we estimated the excess mortality in Thailand from April to October 2021. Our results highlighted that the mortality burden was increased during this period, which would be a major challenge to the healthcare system. Although, Thailand implemented social distancing and stringent lockdown measures, which comprised a national lockdown, border closure, restricting travel, and imposing nighttime curfews. The rapid rise in the number of confirmed cases resulted in the subsequent surge in the number of deaths. Our analysis suggested that death counts directly caused by the COVID-19 infection accounted for approximately 75.0% of the excess deaths. The remaining excess deaths might have been indirectly caused by COVID-19. People, who are ill with other diseases, may struggle to find treatment as the impact of disruptions to the healthcare system during the COVID-19 outbreak (26, 27). There have been reports of inadequate intensive care unit (ICU) beds and ventilators in Thailand (6). A shortage of ICU resources may lead to an increased risk of death for critically ill patients. Some hospitals also lacked blood supplies due to a dramatic decrease in blood donations, resulting in blood shortages throughout the country (28, 29).

Our analysis shows that the excess mortality drastically increases with increasing age, which is broadly consistent with the estimates in other studies (15, 17). The excess mortality was observed mostly in the age group above 60 years of age, accounting for 78.4% of the total excess mortality. These demographic characteristics of mortality agree with the risk of mortality due to the COVID-19 outbreak (30-32). Furthermore, recent clinical studies (33, 34) have found that COVID-19 has a major impact on the elderly. Therefore, targeting primarily the elderly age group (via, such as, vaccination) may reduce hospitalization and mortality rates.

In contrast, the COVID-19 outbreak seemed to have less impact on the younger age group (0–24 years). All-cause mortality in the younger age group may decrease partly due to the reduction in traffic accident mortality. A recent study has also pointed out that the reduction in deaths among the younger age groups may be due to fewer car accidents and work-related accidents during the lockdown period (18). According to the WHO Global Status Report on Road Safety (35), Thailand ranked No.9 in the world and No.1 in Southeast Asia for road traffic deaths, with an estimated road traffic death rate of 33 deaths per 100,000 people in 2018 (36). Moreover, traffic accidents were the third leading cause of death in Thailand in 2020 (37). However, the data in 2021 suggests that traffic accident fatalities have decreased substantially during the course of the COVID-19 outbreak (**Fig. S11** in the supplementary information). The mobility data reported by Apple also suggested that driving and walking mobility tend to decrease during this period. The number of deaths caused by traffic accidents has decreased markedly in the male population (**Fig. S12** in the supplementary information). Specifically, the average number of deaths caused by traffic accidents decreased approximately 20% compared to the previous year. Therefore, our findings might reflect reduced all-cause mortality in the younger age group of men, partly due to the reduction in traffic accident mortality as a result of lockdown measures.

Our results showed higher excess deaths in males than in females. Although the trend of sex-specific deaths in 2021 compared to the previous four years showed a similar trend for men and women, the magnitude of excess mortality is higher for men, specifically in three age groups (25–34, 35–60, and over 60 years of age). There was a noticeable sex difference in excess mortality in age groups older than 25 years. These demographics of excess mortality were also found in other countries such as England and Wales (15), Sweden (17), Italy (38), and Brazil (39). These studies revealed that the majority of excess deaths were attributable to COVID-19. Moreover, other studies suggest that the risk of COVID-related death in males may be related to susceptibility to severe SARS-CoV-2 infections and male hormones (40-42). Therefore, gender could be a risk factor for COVID-related mortality.

The increase in all-cause mortality in Thailand was mainly affected by the COVID-19 outbreak. Besides, during the same COVID-19 outbreak period, Thailand experienced excess pneumonia mortality (**Fig. 6**). Our results showed that pneumonia mortality increased from April to October 2021, corresponding to the period of the COVID-19 outbreak. In this study, we analyzed pneumonia mortality because pneumonia could be a complication of COVID-19 and has symptoms or signs similar to COVID-19 (43, 44). Moreover, among COVID-19 patients, pneumonia was the leading cause of death (45, 46). According to the pneumonia mortality data, the trend of pneumonia deaths continually decreased from 2015 to 2020 (**Fig. S10** in the supplementary information). However, the mortality rate caused by pneumonia increased unexpectedly during the COVID-19 outbreak in April 2021 (22). Furthermore, while the prevalence of influenza during the COVID-19 outbreak in Thailand tended to decrease (47), pneumonia mortality still increased. It is possible that an increase in pneumonia deaths might have been attributed to COVID-19 or misclassified deaths. If an increase in excess pneumonia deaths appears to be misclassification of COVID-19 deaths, this indicates that the true number of COVID-19 deaths might be underestimated. This is a possible explanation for the increase in pneumonia deaths during the COVID-19 outbreak in 2021. However, because there appears to be insufficient evidence and a lack of data during the study period, we were unable to quantify the death toll of misclassified COVID-19 deaths.

Our study, however, has some limitations. In this work, we did not consider other influential factors of death, such as influenza and respiratory syncytial virus infection, due to the lack of data. The empirical findings from N. Suntronwong et al. (47) revealed that the advent of COVID-19 may change human behavior, such as wearing face masks and social distancing, resulting in a decreased transmission of influenza in Thailand. In addition, we did not consider other factors such as effects of heatwaves and ambient air pollution that may correlate with mortality (48, 49) due to the limitation of data. Future work may investigate these factors when the data becomes available.

In summary, our findings showed that the COVID-19 outbreak in Thailand had a huge impact on mortality. During the outbreak in 2021, there was a significant rise in excess fatalities, especially in the older age groups. The increase in mortality was found to be higher in men than in women. It is worth stressing that up-to-date and reliable mortality data are important to assess the impact of the COVID-19 pandemic. Our modeling results could potentially provide insights into the COVID-19 outbreaks and provide a guide for outbreak control and intervention.

## Data Availability

All data produced in the present study are available upon reasonable request to the authors

## Abbreviations

COVID-19: Coronavirus disease 2019
SARS-CoV-2: Severe acute respiratory syndrome - coronavirus - 2
GLMMs: generalized linear mixed models
CI: confidence interval
PI: predictive intervals
AIC: Akaike information criterion

## Acknowledgments

This work was supported by Faculty of Science, Naresuan University. C.M. was supported by the National Research Council of Thailand and the Thailand Center of Excellence in Physics.

## Authors’ Contributions

CW: participated in its design, performed the analysis, wrote the first draft. TL: participated in data extraction and interpretation. CM: participated in its design, analyzed the results, and wrote the manuscript. SC: conceptualized, participated in its design, analyzed the results, and wrote the manuscript. All authors read and approved the final manuscript.

## Availability of Data and Materials

The data supporting the findings can be found in the main paper.

## Declarations

### Ethics approval and consent to participate

All information of COVID-19 and pneumonia surveillance data was collected from the Thai Ministry of Public Health. This study was approved by the Institutional Review Board of Naresuan University (P1-0003/2565) as exemption review. The need of informed consent was waived by the Institutional Review Board of Naresuan University as all data of our study are deidentified.

### Consent for publication

Not applicable

### Author details

^1^Biophysics Group, Department of Physics, Faculty of Science, Mahidol University, Bangkok 10400, Thailand. ^2^Centre of Excellence in Mathematics, CHE, Ministry of Education, Bangkok 10400, Thailand. ^3^Thailand Center of Excellence in Physics, Commission on Higher Education, Bangkok 10400, Thailand. ^4^Department of Physics, Research Center for Academic Excellence in Applied Physics, Faculty of Science, Naresuan University, Phitsanulok 65000, Thailand.

## Supplementary Information

### Deaths due to COVID-19

The number of daily COVID-19 deaths in Thailand (**Fig S1**) was retrieved from the Center for Systems Science and Engineering (CSSE) at Johns Hopkins University (1).

**Figure S1.**
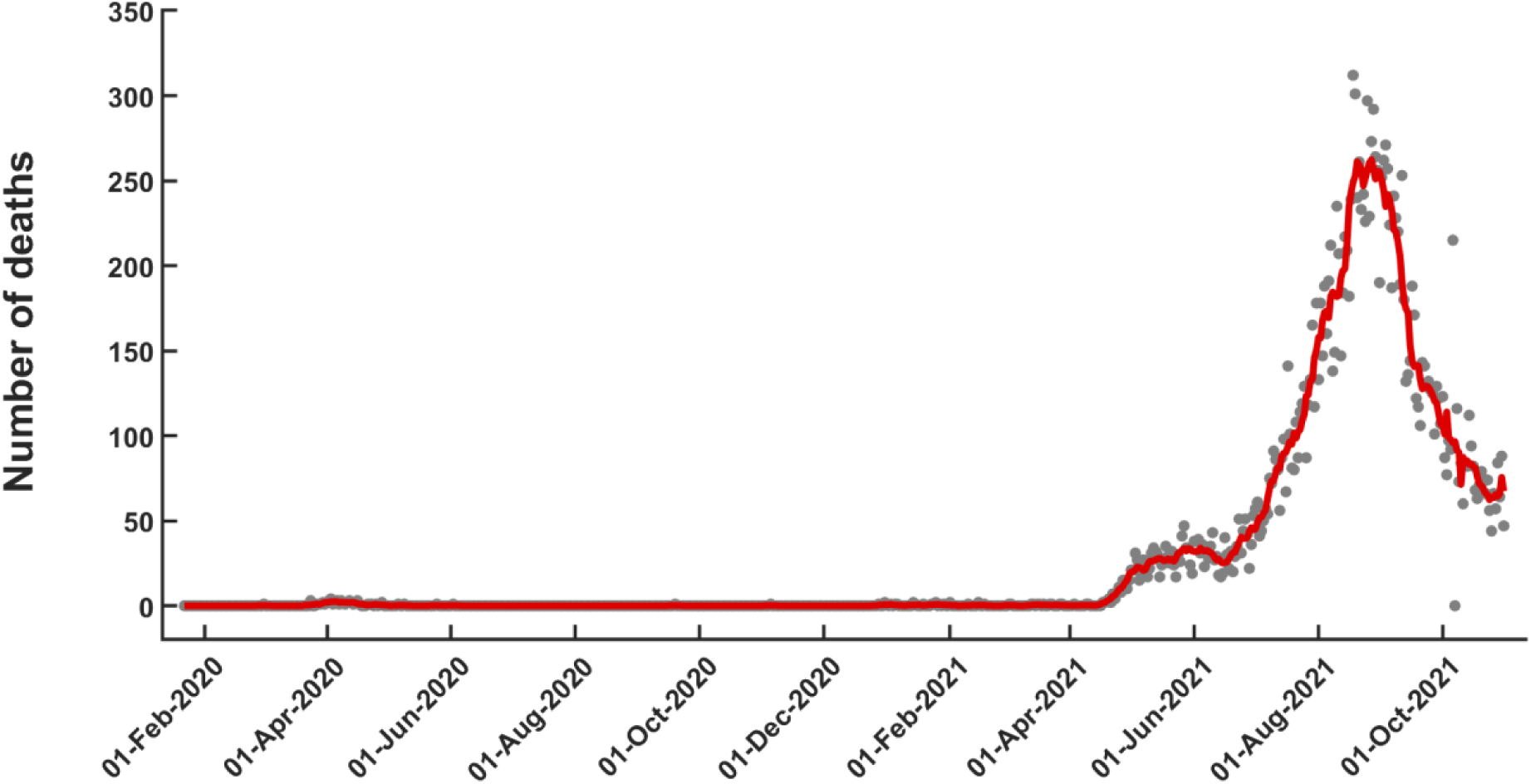
Daily confirmed COVID-19 deaths. The red line illustrates the 7-day average of the number of COVID-19 deaths starting from January 22, 2020, to October 31, 2021 in Thailand. Points show the observed daily mortality data.

### Baseline mortality during 2015 – 2021

We used mortality data from January 2015 to December 2019 by age and sex to fit the models. For Model A, the structure of the model was described in the main text. The observed and baseline mortality of Model A stratified by age group in 2015–2021 for men and women are shown in **Fig. S2** and **S3**, respectively.

**Figure S2.**
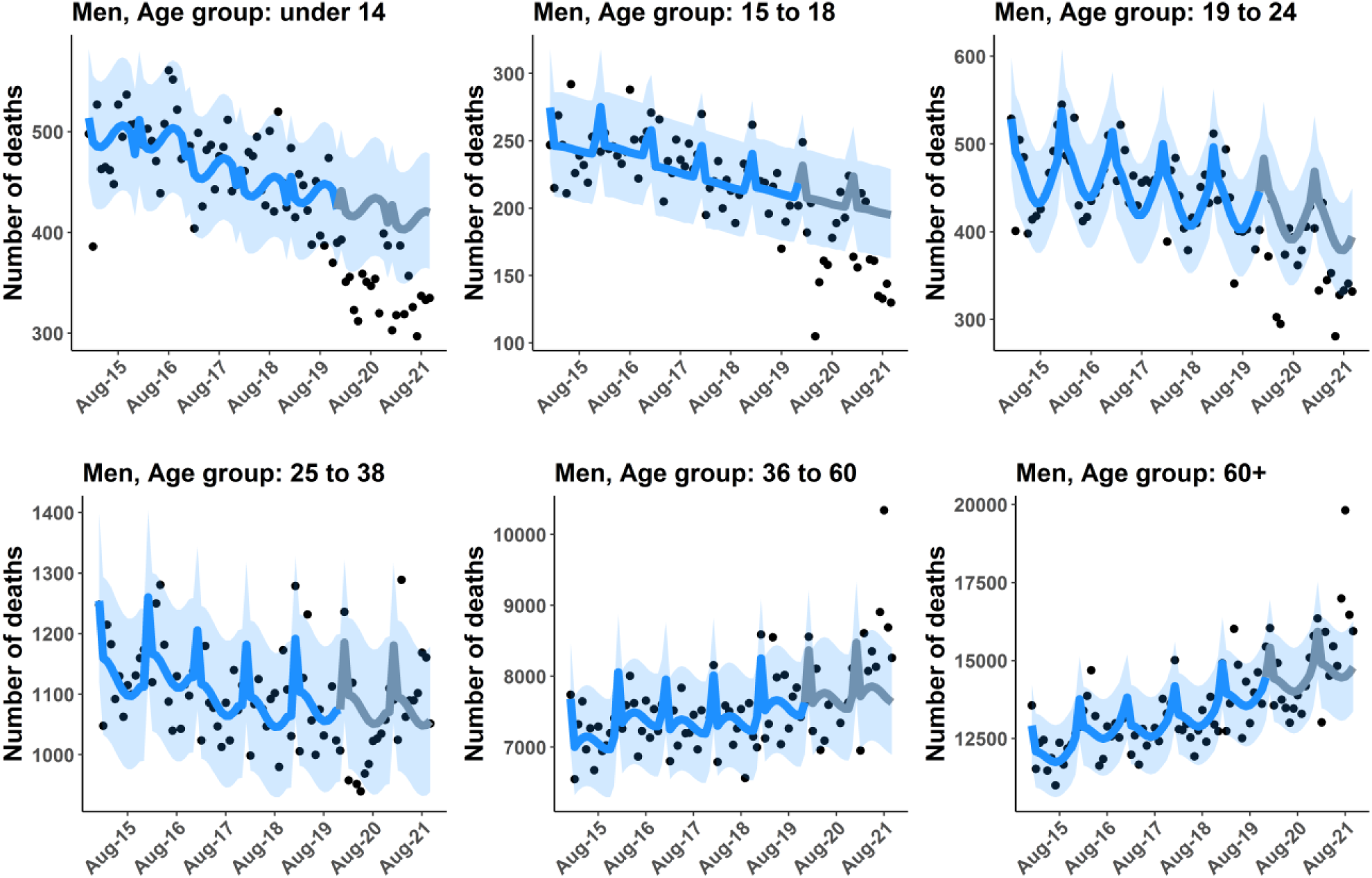
Men baseline mortality from Model A. Lines illustrate the baseline mortality by six age groups (0-14, 15-18, 19-24, 25-34, 35-60, and over 60 years of age). Grey lines show the predicted baseline starting from January 2020 to October 2021. Points show the observed mortality data. Shaded areas indicate 95% CI.

**Figure S3.**
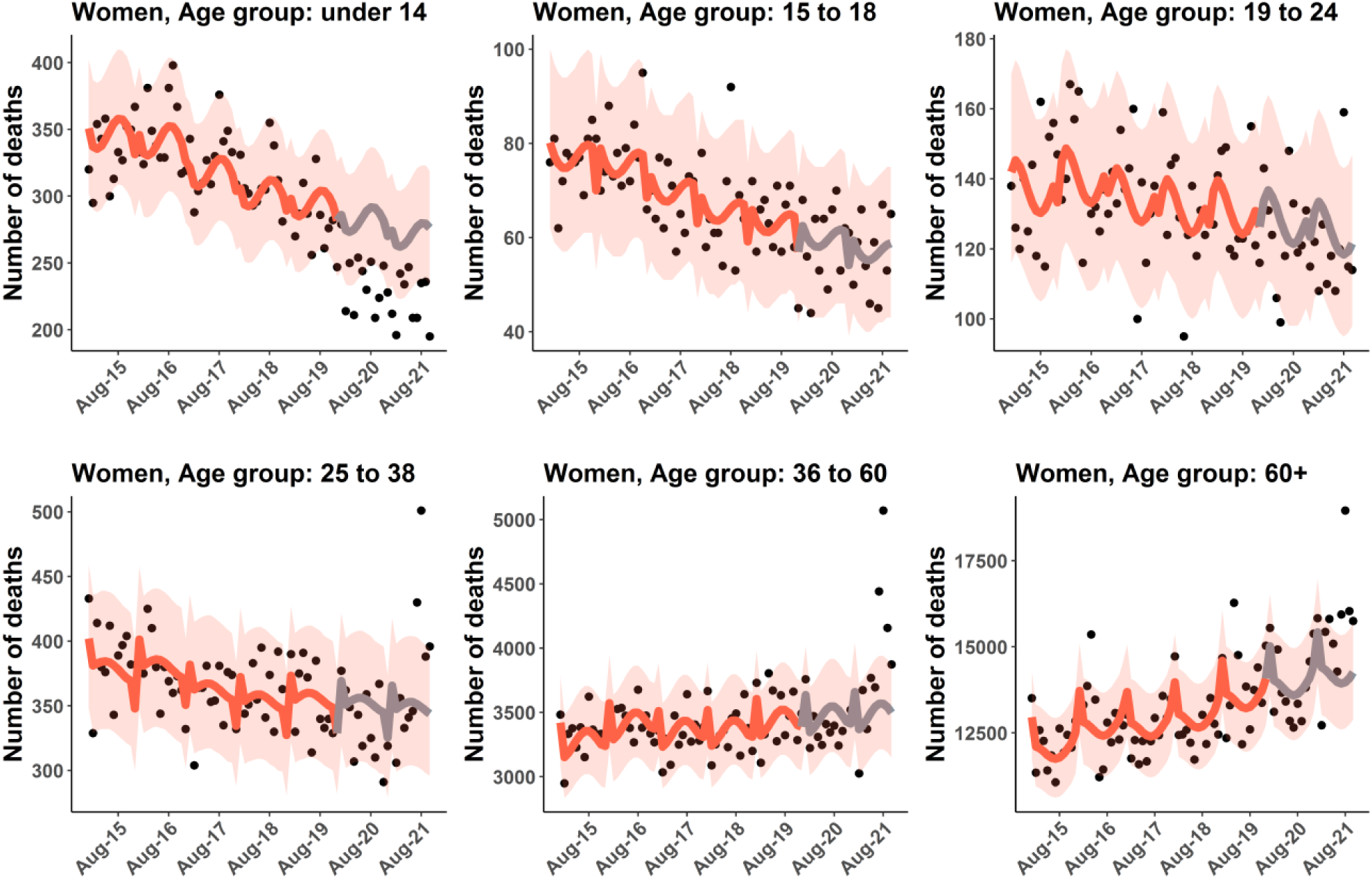
Women baseline mortality from Model A. Lines illustrate the baseline mortality by six age groups (0-14, 15-18, 19-24, 25-34, 35-60, and over 60 years of age). Grey lines show the predicted baseline starting from January 2020 to October 2021. Points show the observed mortality data. Shaded areas indicate 95% CI.

The other three models have different structures, as follows.

#### Model B

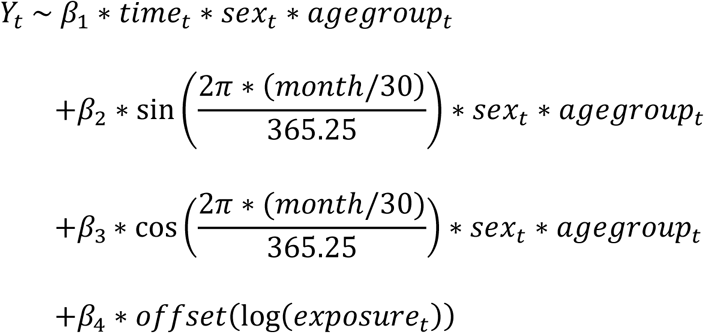

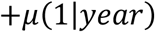

The observed and baseline mortality stratified by age group in 2015-2021 for men and women, are shown in **Fig S4-S5**, respectively.

**Figure S4.**
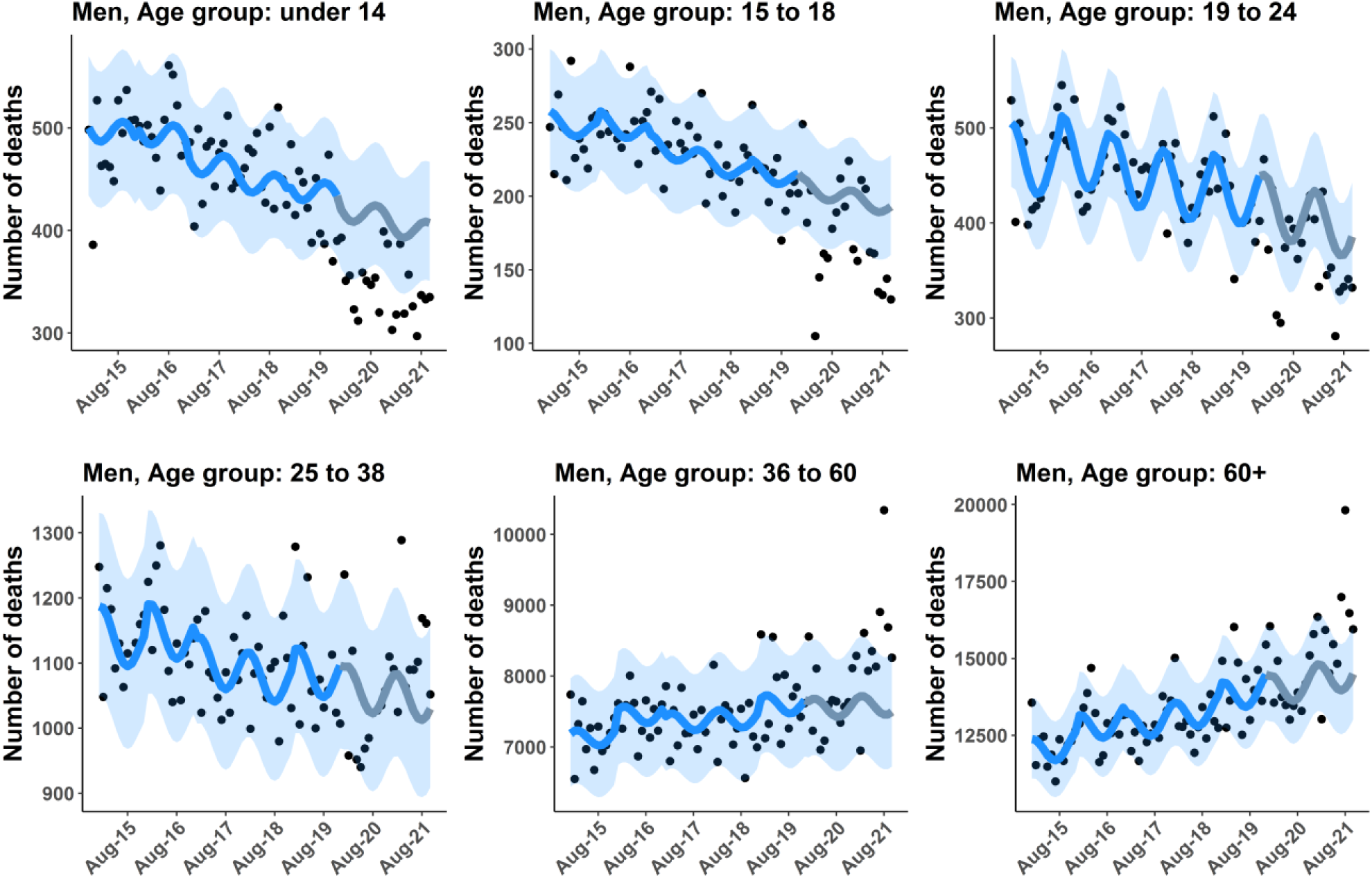
Men baseline mortality from Model B. Lines illustrate the baseline mortality by six age groups (0-14, 15-18, 19-24, 25-34, 35-60, and over 60 years of age). Grey lines show the predicted baseline starting from January 2020 to October 2021. Points show the observed mortality data. Shaded areas indicate 95% CI.

**Figure S5.**
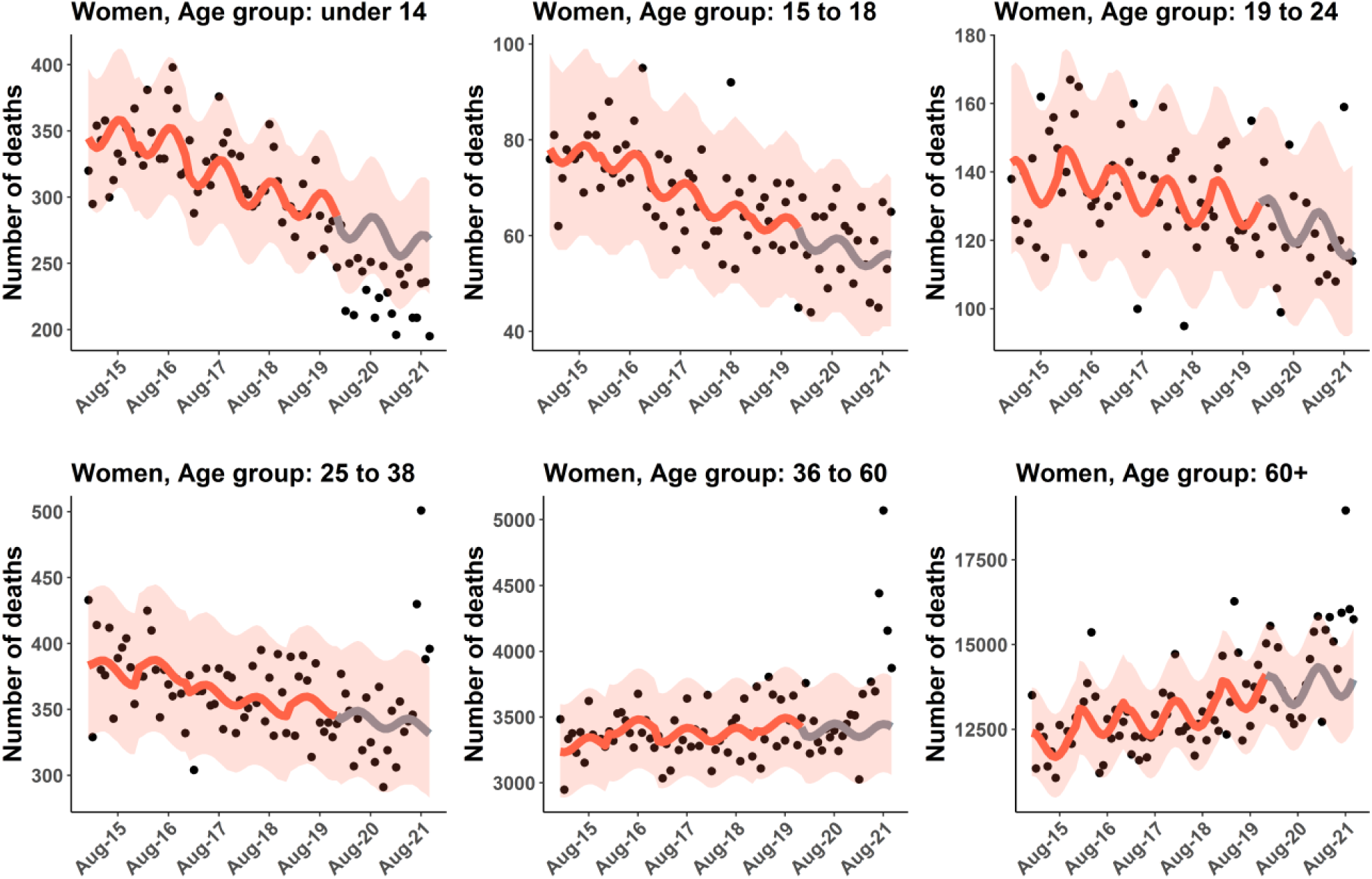
Women baseline mortality from Model B. Lines illustrate the baseline mortality by six age groups (0-14, 15-18, 19-24, 25-34, 35-60, and over 60 years of age). Grey lines show the predicted baseline starting from January 2020 to October 2021. Points show the observed mortality data. Shaded areas indicate 95% CI.

#### Model C

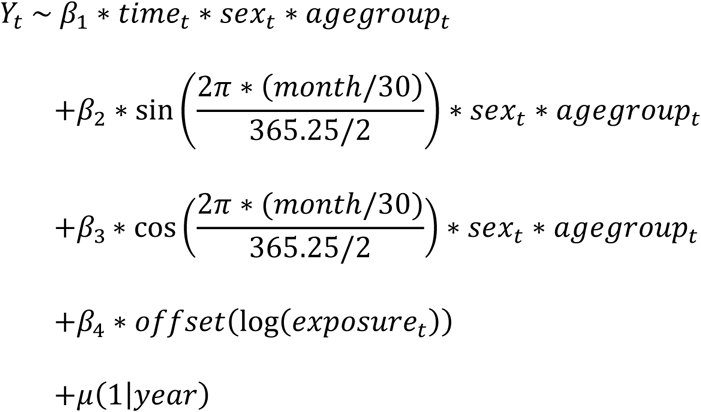

The observed and baseline mortality stratified by age group in 2015-2021 for men and women, are shown in **Fig S6-S7**, respectively.

**Figure S6.**
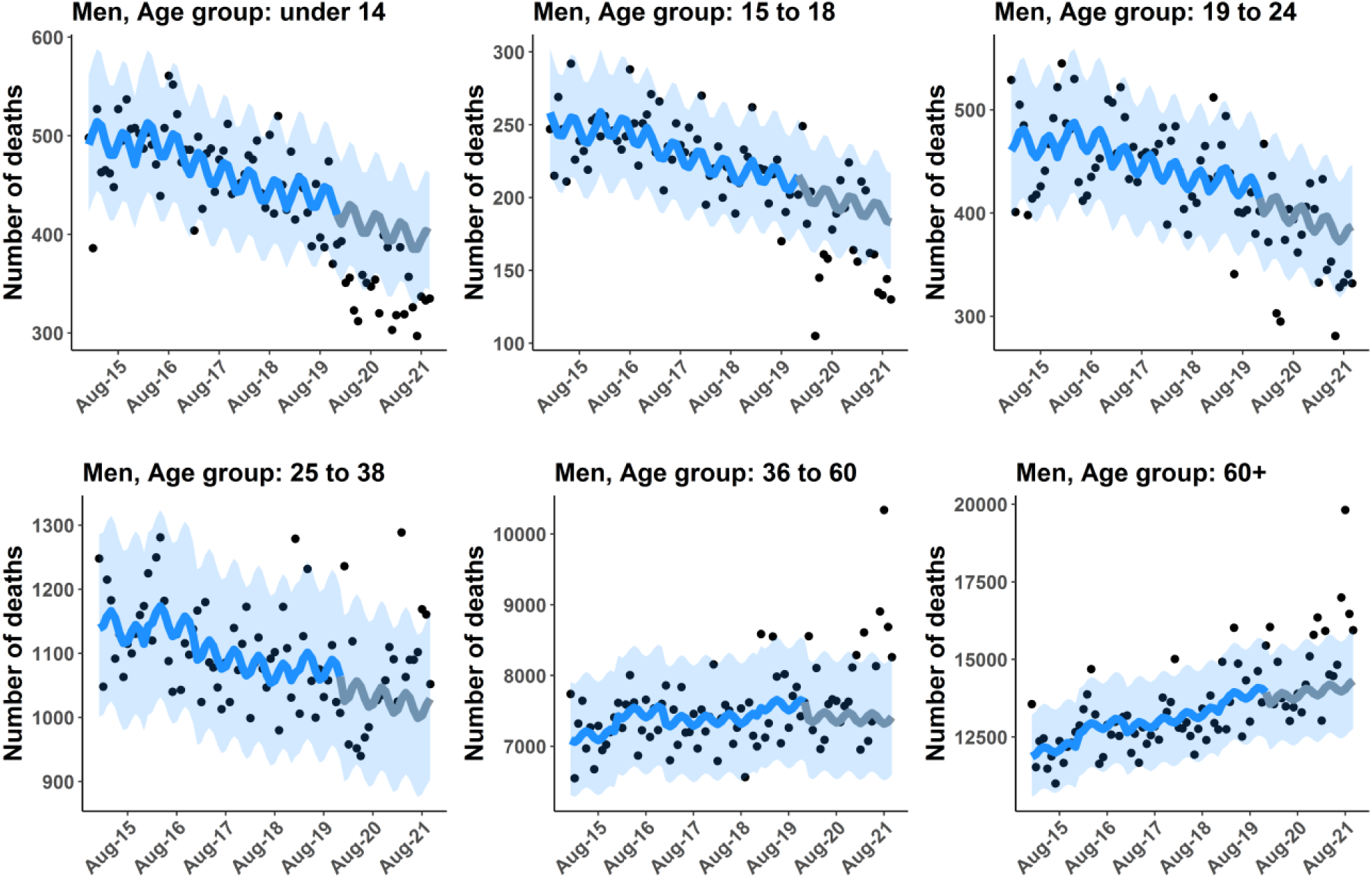
Men baseline mortality from Model C. Lines illustrate the baseline mortality by six age groups (0-14, 15-18, 19-24, 25-34, 35-60, and over 60 years of age). Grey lines show the predicted baseline starting from January 2020 to October 2021. Points show the observed mortality data. Shaded areas indicate 95% CI.

**Figure S7.**
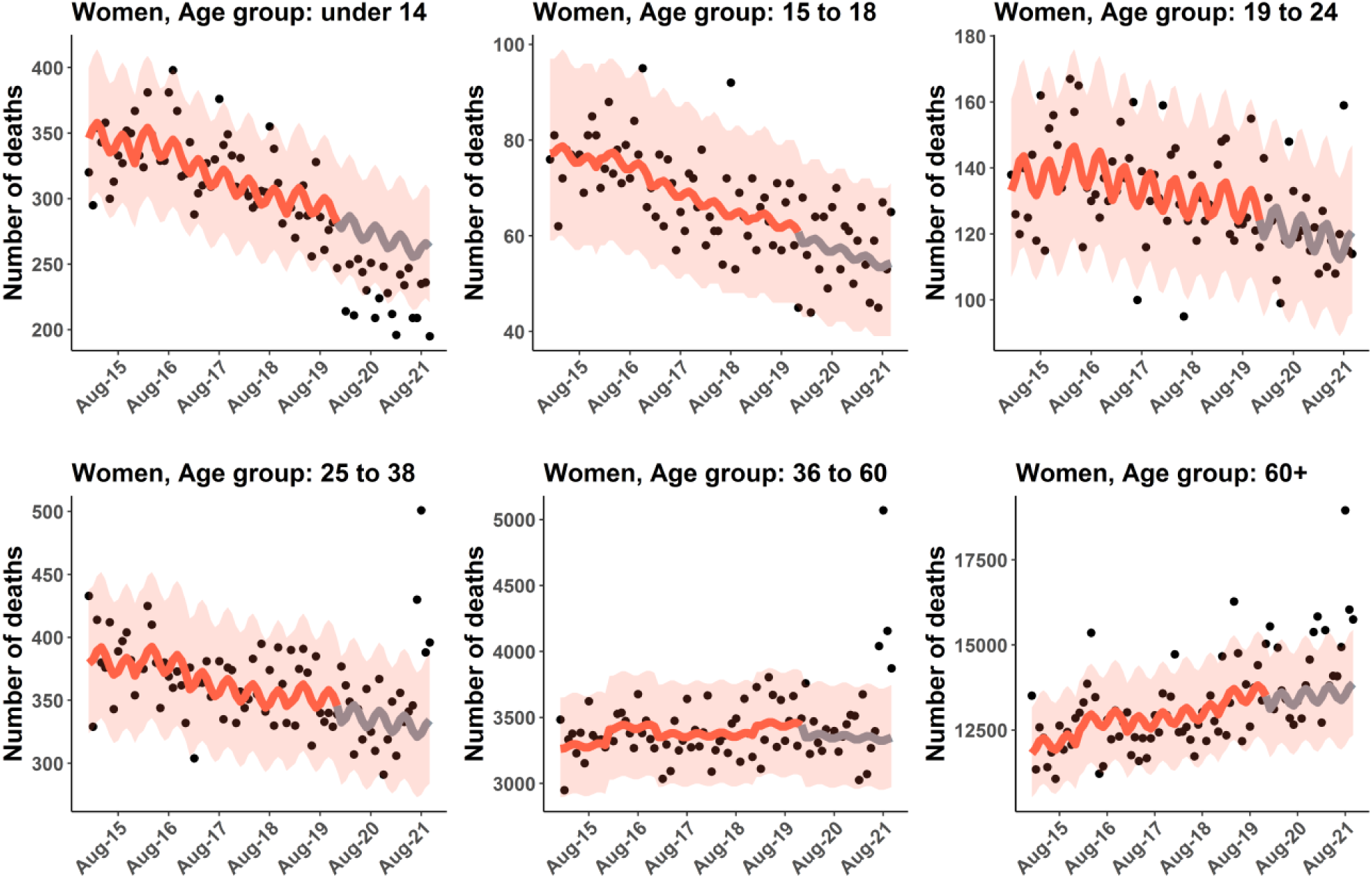
Women baseline mortality from Model C. Lines illustrate the baseline mortality by six age groups (0-14, 15-18, 19-24, 25-34, 35-60, and over 60 years of age). Grey lines show the predicted baseline starting from January 2020 to October 2021. Points show the observed mortality data. Shaded areas indicate 95% CI.

#### Model D

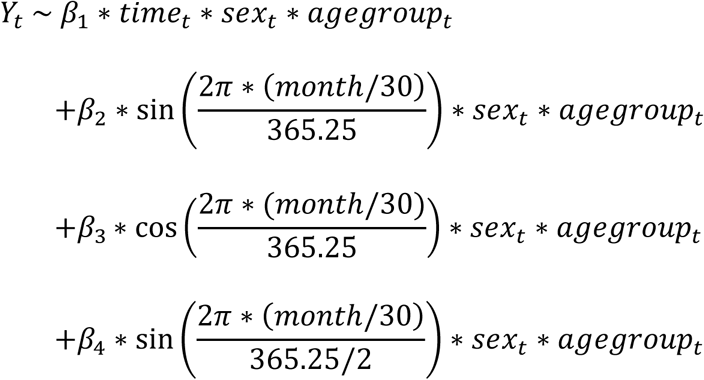

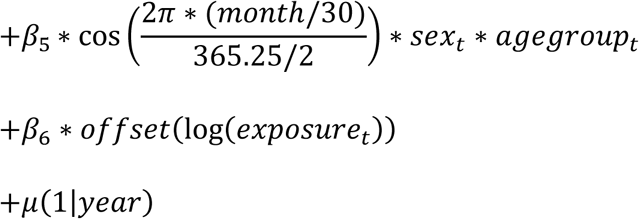

**Figure S8.**
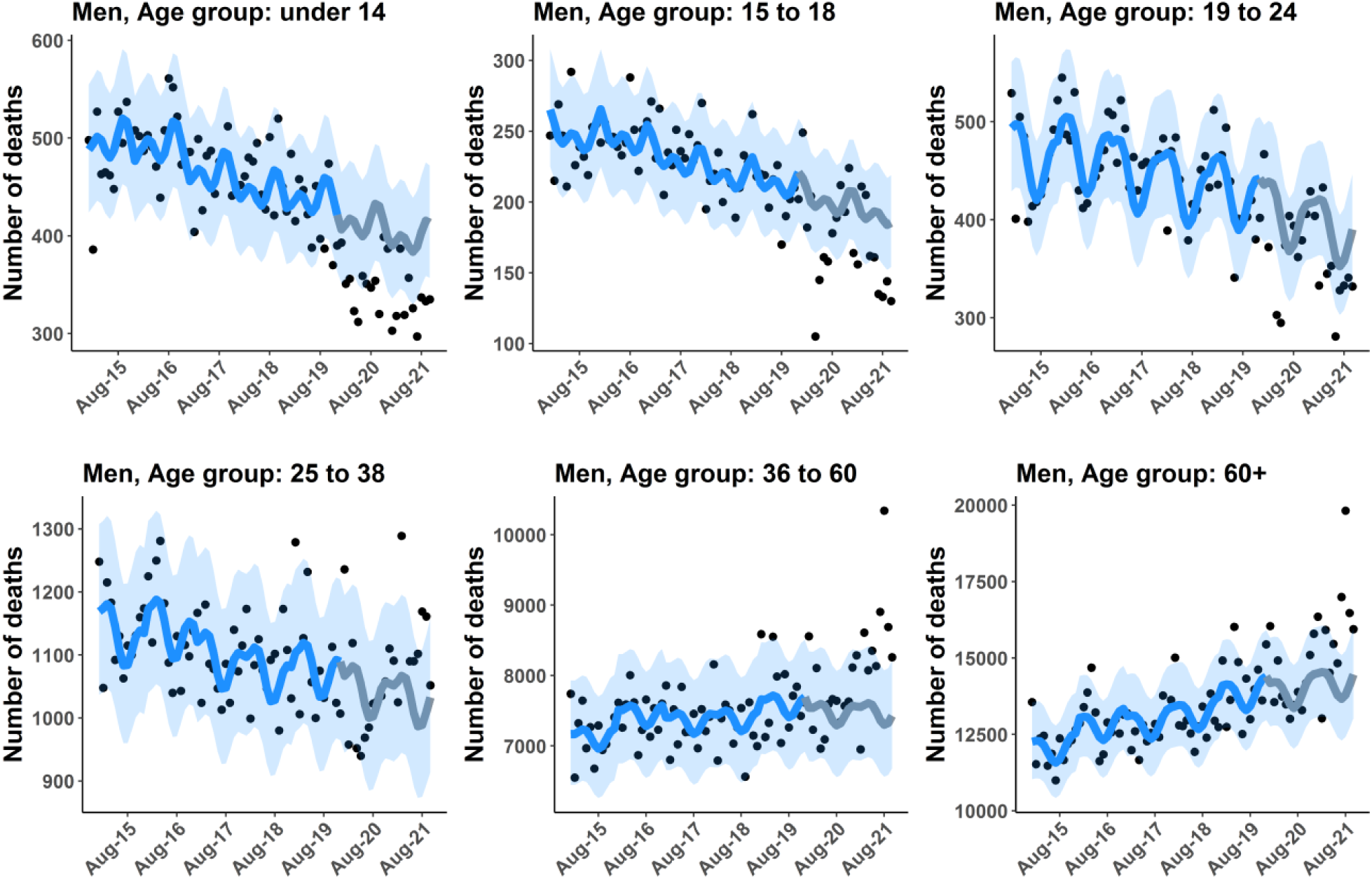
Men baseline mortality from Model D. Lines illustrate the baseline mortality by six age groups (0-14, 15-18, 19-24, 25-34, 35-60, and over 60 years of age). Grey lines show the predicted baseline starting from January 2020 to October 2021. Points show the observed mortality data. Shaded areas indicate 95% CI.

**Figure S9.**
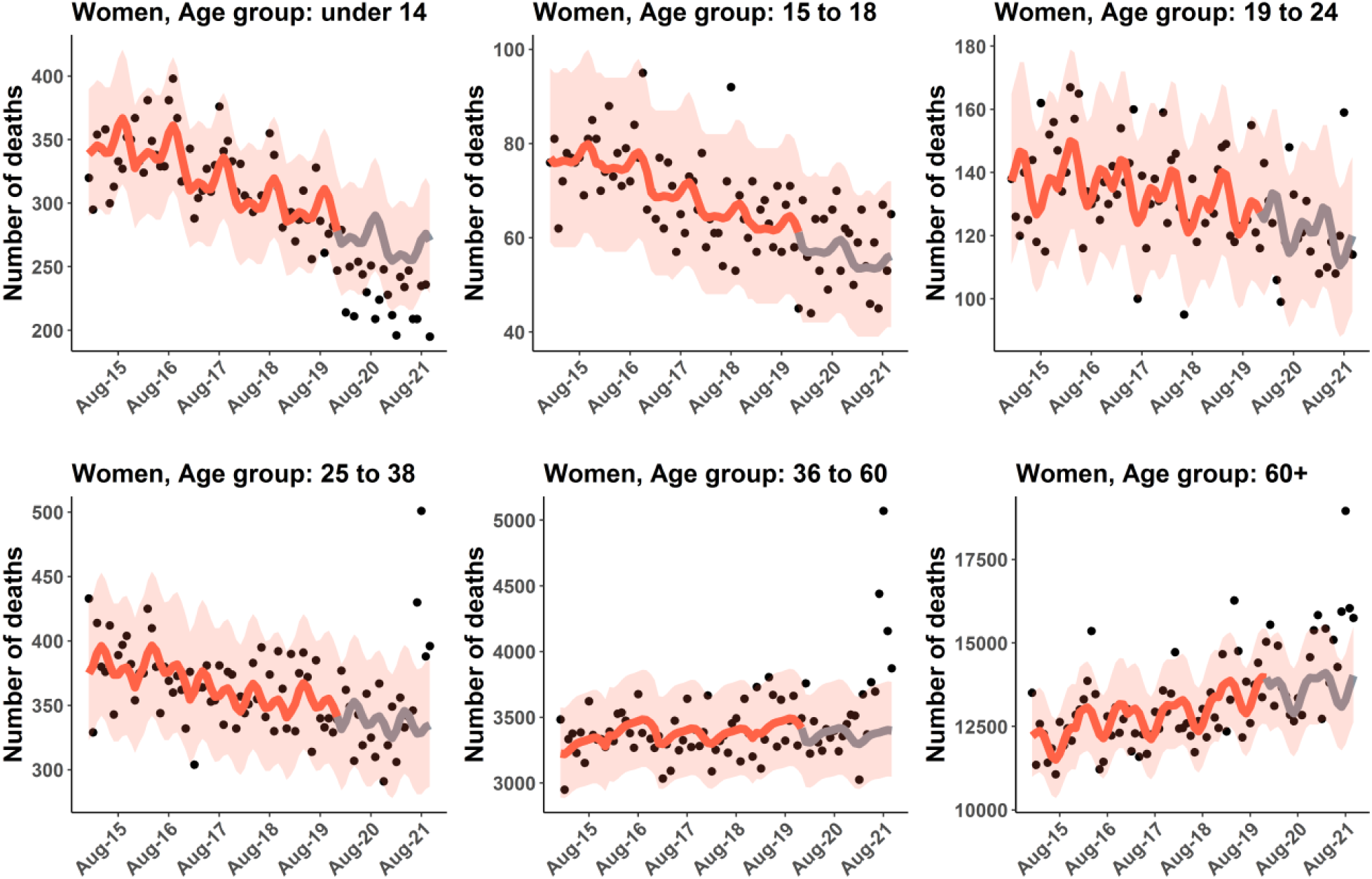
Women baseline mortality from Model D. Lines illustrate the baseline mortality by six age groups (0-14, 15-18, 19-24, 25-34, 35-60, and over 60 years of age). Grey lines show the predicted baseline starting from January 2020 to October 2021. Points show the observed mortality data. Shaded areas indicate 95% CI.

**Table S1.**
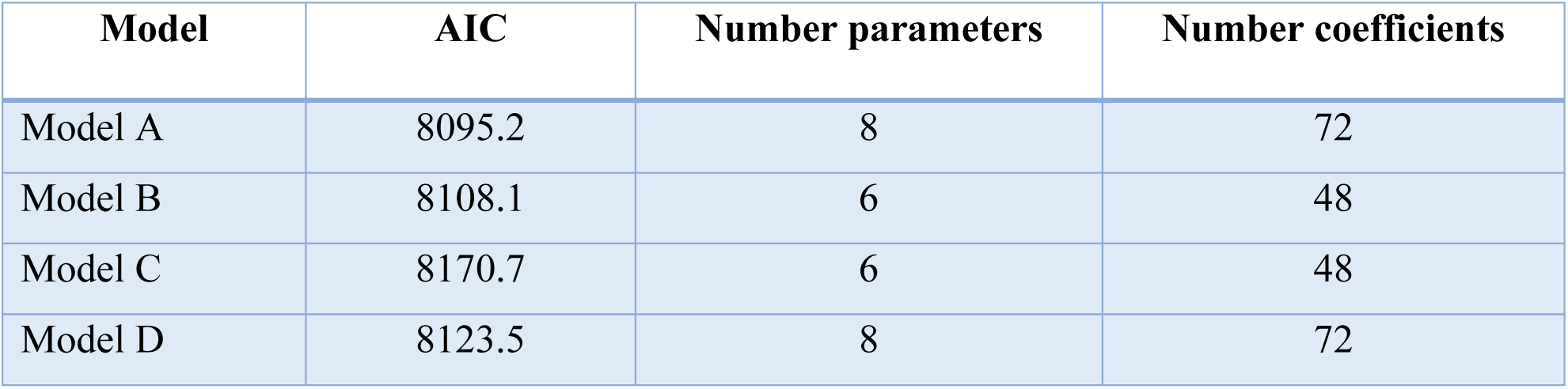
Model comparison.

The observed and baseline mortality stratified by age group in 2015-2021 for men and women, are shown in **Fig S8-S9**, respectively.

A summary of four different models is shown in **Table S1**. We used AIC to estimate the quality of each model.

### Pneumonia excess mortality

We used the pneumonia mortality data from January 2015 to December 2019 to estimate the baseline pneumonia mortality in the absence of COVID-19. The monthly pneumonia mortality data in Thailand was obtained from the Bureau of Epidemiology, Department of Disease Control, MoPH, Thailand (2). The mortality data was fitted by generalized linear mixed models (GLMMs) as follows:

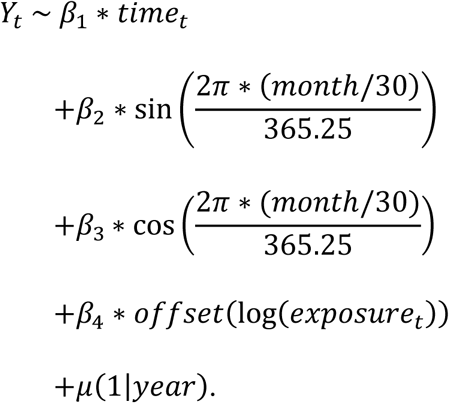

We then projected the pneumonia baseline mortality forward from January 2020 to October 2021. The observed and baseline mortality of pneumonia in Thailand is illustrated in **Fig S10**. The pneumonia excess mortality was then calculated from the number of pneumonia deaths minus the baseline prediction.

**Figure S10.**
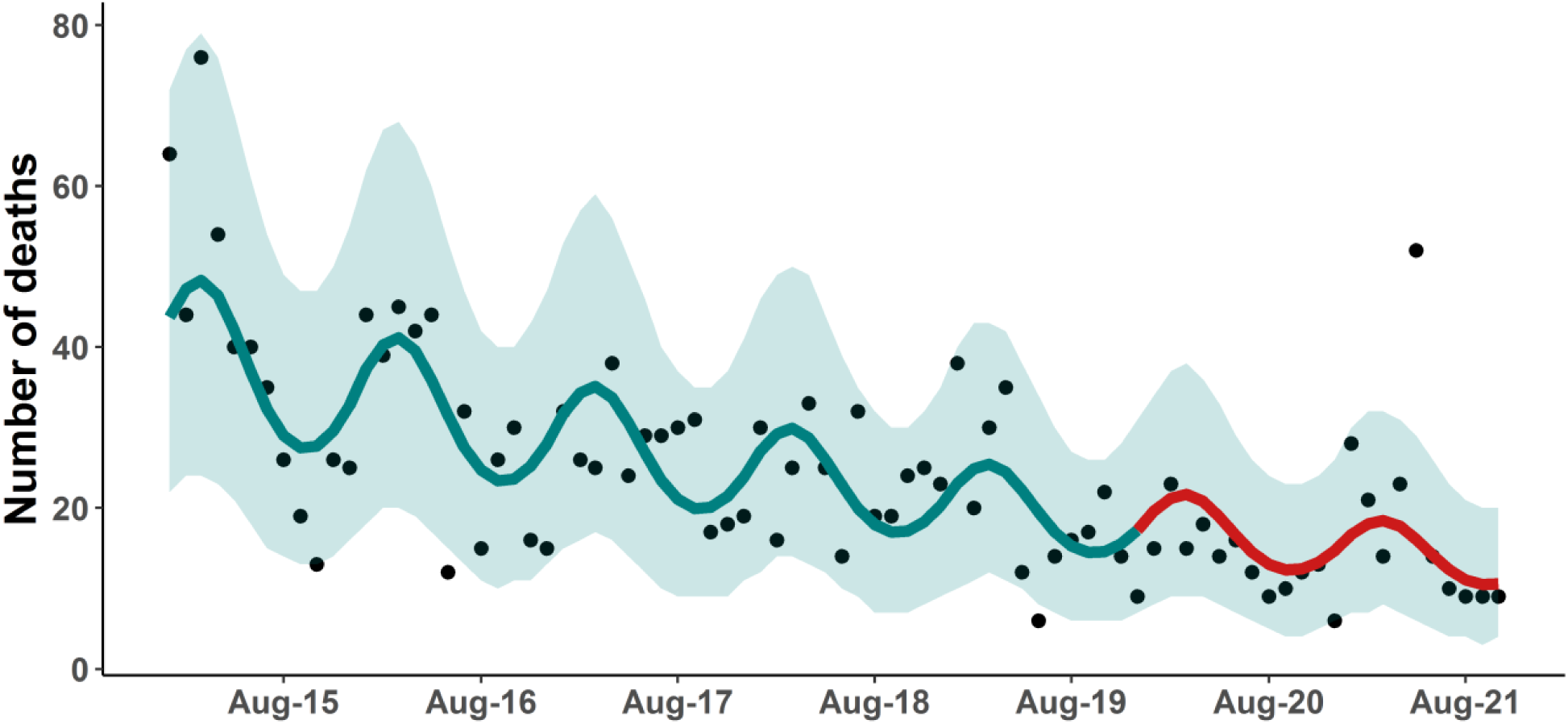
Pneumonia baseline mortality. Line illustrates the pneumonia baseline mortality in Thailand starting from January 2015 to October 2021. The red line shows the predicted pneumonia baseline mortality starting from January 2020 to October 2021. Points show the observed pneumonia mortality data. The shaded area indicates 95% CI.

### Traffic accident mortality data

**Figure S11** shows the traffic accident mortality deaths from January 2017 to October 2021 and the mobility trends in Thailand. Data on traffic accident-related deaths was acquired from the Road Accidents Data Center for Road Safety Culture of Thailand (Thai RSC) (3). The accident mortality data were also stratified by age group and gender (**Fig S12**). The number of people dying from traffic accidents was categorized into six age groups (0-14, 15-18, 19-24, 25-34, 35-60, and over 60 years of age). The data suggested that after the Thai government implemented social distancing and stringent lockdown measures, the number of traffic accident deaths decreased by approximately 20% compared with the average traffic accident deaths of the previous year. Moreover, we used the mobility in Thailand reported by Apple (4) as a representative of the mobility in Thailand during the lockdown. Driving and walking mobility tend to decrease compared with baseline.

**Figure S11.**
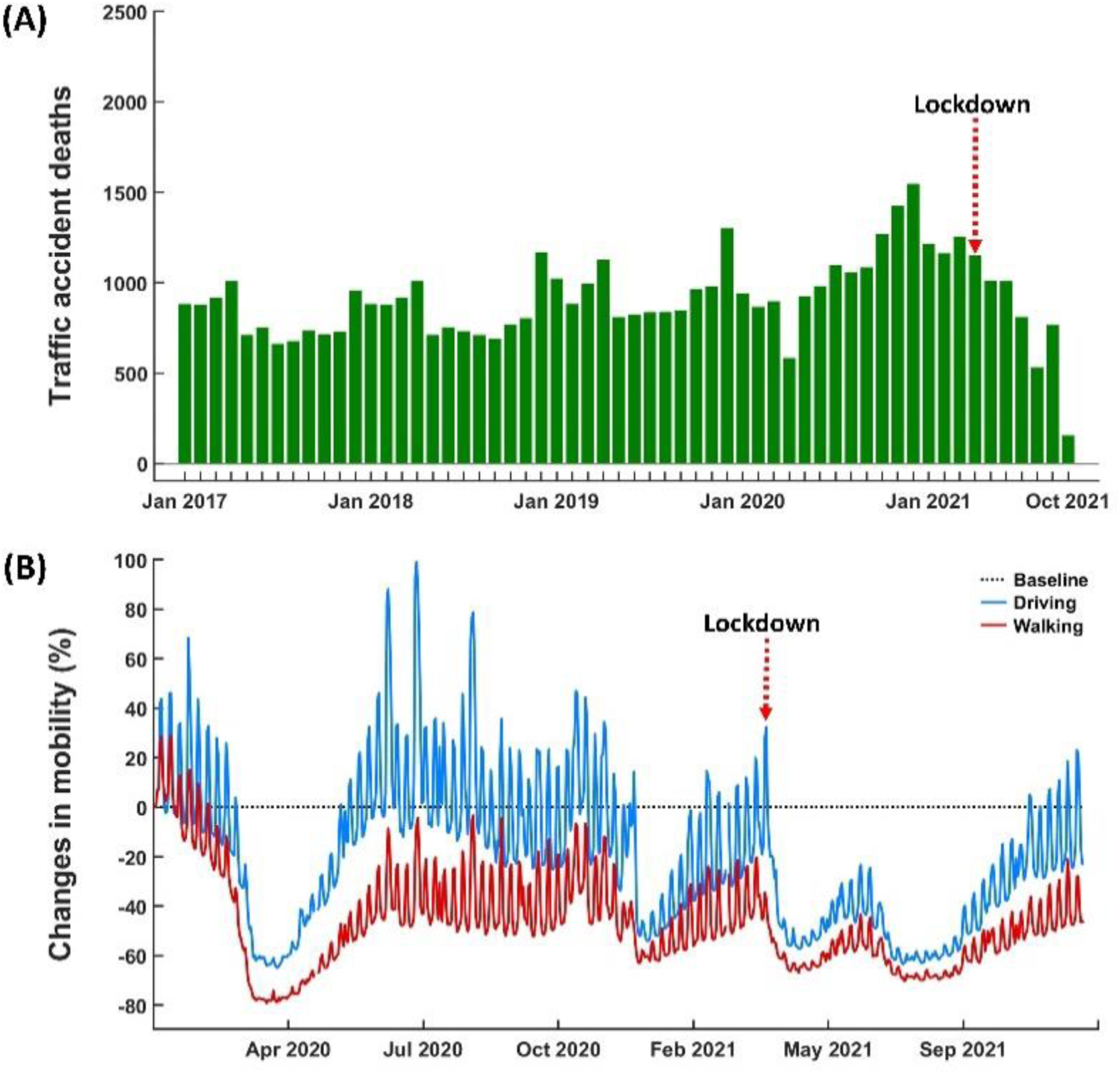
Traffic accident mortality data and mobility trends data. **(A)** Traffic accident mortality data from January 2017 to October 2021. **(B)** The mobility in Thailand reported by Apple.

**Figure S12.**
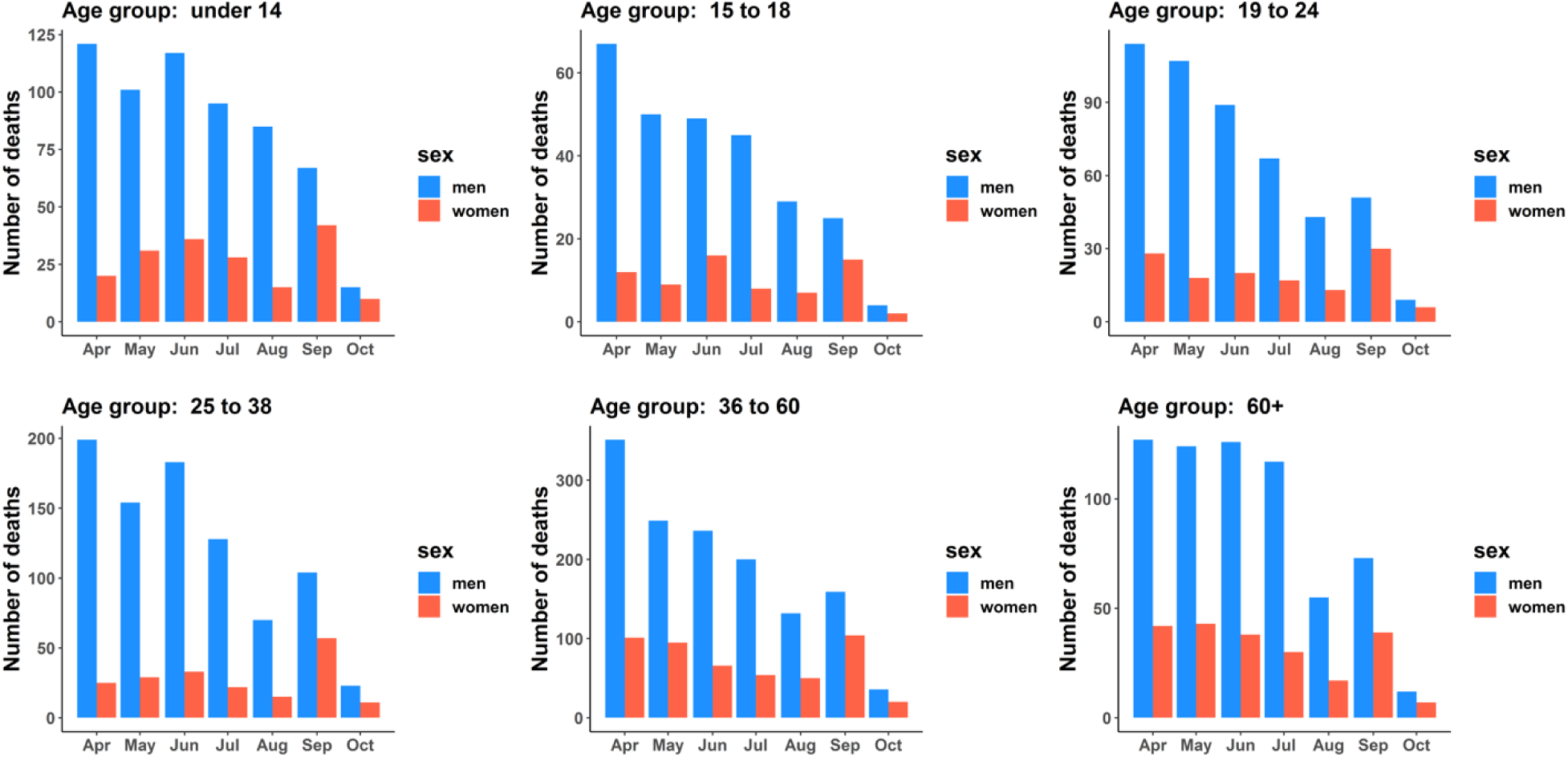
Traffic accident deaths by gender and age groups. Bars show the monthly traffic accident deaths by gender and age groups starting from April to October 2021.

